# A REGISTER-BASED, EPIDEMIOLOGICAL EVALUATION OF DISEASE ACTIVITY AND FUNCTIONAL PERFORMANCE IN ANKYLOSING SPONDYLITIS PATIENTS AFTER MULTIMODAL SPA THERAPY IN AUSTRIA

**DOI:** 10.1101/2025.08.04.25332936

**Authors:** Kathrin Maria Bogensberger, Sonja Wildburger, Julia Fuchs, Bertram Hoelzl, Rudolf Radlmueller, Wolfgang Foisner, Heiko Rychlowski, Martin Offenbaecher, Martin Gaisberger, Markus Ritter, Antje van der Zee-Neuen

**Author notes:** **<u>Correspondence to:</u>** PD. Dr. Antje van der Zee-Neuen, Paracelsus Medical University, Strubergasse 22, A-5020 Salzburg, AUSTRIA, /.

## Abstract

**Objective:** Evidence regarding the impact of multimodal treatment on disease activity and functional performance in ankylosing spondylitis (AS) remains scarce. This study aims to evaluate changes in these outcomes after multimodal rehabilitation, using the Bath Ankylosing Spondylitis Disease Activity Index (BASDAI) and Functional Index (BASFI).

**Methods:** Mixed-effects linear regression was performed on prospectively collected registry data to explore associations between timepoints and BASDAI/BASFI. Predicted linear margins from fixed effects were calculated for each timepoint. Data were derived from AS patients who underwent multimodal rehabilitation and completed questionnaires at baseline (T0), post-intervention (T1), and at 3 (T2), 6 (T3), and 9 (T4) months follow-up.

**Results:** Twohundred-fourtynine AS patients were included (52.14 ± 10.20 years; 42.57% female, BMI: 26.64 ± 4.27 kg/m2). Compared to baseline, BASFI/BASDAI improved significantly until 6 and 9 months, respectively. Predicted BASDAI scores at T0, T1, T2 and T3 were 2.99 (95%CI 2.77; 3.21), 2.31 (2.08; 2.53), 2.38 (2.15; 2.61) and 2.69 (2.46; 2.92). At T4 the score was higher than at T0 (3.04 (2.81; 3.28)). Compared to T0, BASDAI score at T1 reflected a clinically relevant change. Predicted BASFI scores at T0, T1, T2, T3 and T4 were 4.09 (3.88; 4.31), 2.71 (2.49; 2.94), 2.83 (2.60; 3.06), 3.24 (3.01; 3.46) and 3.59 (3.36; 3.82). Compared to T0, BASFI scores at T1, T2 and T3 reflected a clinically relevant change.

**Conclusion:** Significant improvements in physical functioning and disease activity among AS patients following multimodal rehabilitation were found. Such interventions may serve as a valuable complement to pharmacological treatment.

**What is really known on this topic:**

- Non-pharmacological, multimodal treatment approaches play a key role in treating AS patients by complementing pharmacological treatment regimens.

**What this study adds:**

- To our knowledge this is the first study to evaluate changes in BASFI and BASDAI in context of multimodal spa therapy.
- Significant improvements in both BASFI and BASDAI up to 9 months after treatment.

**How this might affect research, practice, or policy:**

- The findings may complement the lack of knowledge in context of disease activity and physical performance after multimodal treatment interventions.
- The study might also emphasize the determination of adequate treatment intervals.

## Introduction

Ankylosing spondylitis (AS) is a form of spondyloarthropathy (SpA) and is defined as a chronic, inflammatory and autoimmune rheumatic disease of unknown etiology. AS predominantly affects the articulations of the spine, the joints linking the pelvis and lower spine (i.e. sacroiliac joints) as well as adjacent tendons and ligaments [1]. Patients with AS may experience extra-articular effects, including acute anterior uveitis and inflammatory bowel disease. Such extra-articular manifestations occur in approximately 5-10% of AS patients in Western countries [2, 3]. AS is characterized by severe pain and inflammation, impairments in functional performance as well as axial deformity affecting patients’ participation in activities of daily living and quality of life (QoL) [4, 5].

In order to maintain spinal flexibility, reduce the impact of symptom burden and functional limitations, and improve QoL, a variety of treatment strategies are available. These encompass pharmacological and non-pharmacological treatments as well as surgical procedures [1, 6]. In terms of pharmacological treatments, nonsteroidal anti-inflammatory drugs (NSAIDs) and TNF-α inhibitors (TNFis) have been identified as the preferred option. In addition, further medication options include non-TNFi biologics, methotrexate, sulfasalazine, and JAK inhibitors [1, 7]. Concomitant non-pharmacological, multimodal treatments have demonstrated effectiveness in the management of AS, irrespective of disease severity and pharmacological management. The term ‘multimodal’ refers to treatment methods that are not limited to exercise regimens, but also include educational, alternative and complementary interventions [6, 8, 9]. The purpose of this more holistic approach is to enhance self-management, motivation and compliance, but also to provide symptom relief, particularly regarding QoL and symptom-related outcomes including physical performance, workability, and psychological health [10]. One potential component of such a complementary approach is hyperthermia treatment. This is defined as the application of thermal energy in different forms for therapeutic purposes, such as balneotherapy [11, 12]. Despite the heterogeneity of evidence, hyperthermia treatment has shown beneficial effects on QoL, overall well-being, and pain [12–15].

In accordance with the ASAS/EULAR recommendations, the primary objective of disease management is the optimization of long-term QoL. This is to be achieved by means of controlling symptom burden and inflammation, preventing progressive structural damage, preserving or normalizing function, and facilitating social participation. Disease domains that have been identified as contributing to QoL include, among others, disease activity and functional performance [6]. To achieve a more comprehensive assessment, it is imperative that patient-reported outcomes (PROs), in addition to clinical findings, laboratory tests and imaging, are incorporated into the overall disease monitoring [6, 10, 16]. In the context of disease management of AS, the Bath Ankylosing Spondylitis Disease Activity Index (BASDAI) and the Bath Ankylosing Spondylitis Functional Index (BASFI) are effective PROs for the monitoring and evaluation of AS treatment strategies [6]. The present study sought to explore the course of BASFI and BASDAI in individuals with AS after undergoing multimodal spa therapy incorporating low-dose radon. It was hypothesized that both outcomes would improve substantially after treatment.

## Methods

A longitudinal analysis was conducted using prospectively collected register data from the ongoing “Radon indication registry for the assessment of pain reduction, increase in quality of life, and improvement in body functionality throughout low-dose radon hyperthermia therapy.” This registry, identified by the registration ID ISRCTN67336967 (https://doi.org/10.1186/ISRCTN67336967), is based in the Gastein valley in Austria.

### Intervention

Patients included in the registry receive a comprehensive multimodal, non-pharmacological spa therapy in the valley of Gastein, embedded in the Austrian Alps. The therapy encompasses a range of treatments prescribed by local physicians, with specific focus on low-dose radon treatment. The typical intervention program comprises a diverse and individualized set of therapeutic modalities, such as physical exercise, massages, lymphatic drainage, mud therapy, ergometry, progressive muscle relaxation, trainings and consultations for back pain prevention, anti-smoking guidance, and nutrition education. Additionally, patients undergo medical examinations and may attend informative lectures. Importantly, all participants receive low-dose radon balneotherapy and/or low-dose radon speleotherapy. The balneotherapeutic radon treatment involves immersing patients in thermal water at approximately 37°C, infused with low levels of radon (averaging 707.57 Bq/L, measured by liquid scintillation using Triathler™ LSC Hidex). The thermal water contains essential elements in specific concentrations (mg/kg water): Na^+^ 80.01; K^+^ 5.71; Li^+^ 0.27; Ca^2+^ 19.84; Mg^2+^ 0.75; HCO_3_^−^ 57.91; Cl^−^ 24.96; F^−^ 5.61; SO_4_^2−^ 130.67; H_2_SiO_3_ dissociated 46.16/colloidal 8.17; H_3_BO_3_ 0.40; CO_2_ 6.87. Trace elements are present as follows (mg/L water): Hg 0.0008; As <0.02; Pb 0.08; Cr <0.01; Se <0.05; Cd <0.01; CN^−^ <0.01. Typically, patients undergo around 10 balneotherapeutic radon baths, each lasting 20 minutes. The speleotherapeutic radon treatment involves relaxation in the Gastein healing gallery, a former mine originally excavated in search of gold. Patients spend an average of 60 minutes in this gallery on alternate days, amounting to an average of 11 speleotherapy sessions. The gallery is situated at a moderate altitude (1270 m above sea level) and features a low atmospheric radon activity (averaging 44 kBq/m3 air, as determined by the healing gallery), high humidity (70-100%), and ambient temperatures ranging from 37 to 41.5°C.

The total therapy duration averages 17.5 days, with a standard deviation of 3.5 days. Of note: the specific combination of treatments may vary for each patient, and the study does not distinguish between these combinations due to limited information on the exact combination of treatment components for each patient at the time of data analysis. A potential intervention might include 1 individual consultation for back pain prevention, participation in a nutrition advice group, 14 group exercise therapy sessions, 8 massage therapy sessions, 6 mudpacks, 6 balneotherapeutic radon applications, 3 group consultations for back pain prevention, 8 underwater massages, and 11 hydrotherapeutic exercise sessions. Additional or alternative sessions or therapeutic approaches are determined at the discretion of the spa physicians responsible.

### Patient recruitment

Patients diagnosed with a rheumatic musculoskeletal disorder (RMD) before their visit to the Gastein valley for spa therapy are eligible for inclusion in the radon registry and are recruited by physicians from participating spa centers. Upon obtaining informed consent, patients complete standardized paper questionnaires at multiple time points, including just before (baseline), immediately after, and at 3, 6, and 9 months following the multimodal spa therapy, which includes low-dose radon treatment. These questionnaires gather essential data on sociodemographic characteristics, pain levels, quality of life, functional ability, and disease activity. The first two questionnaires are handed over in person by medical staff, while the remaining three questionnaires are sent out to patients. Patients return the completed questionnaires to the spa centers via provided return envelopes. To ensure privacy, personal information that could identify participants is removed by the spa facility’s medical staff, and pseudonymized data is then transferred to the employees of the Gastein Research Institute. There, a research assistant manually enters the data into a digital database for analysis. As for the analysis for this manuscript, the radon indication registry included a total of 249 subjects with AS who had completed questionnaires at all specified time points with no missing values in any of the variables considered for analysis.

### Outcomes, main independent variable of interest and covariates

The German version of the Bath Ankylosing Spondylitis Disease Activity Index (BASDAI) and the Bath Ankylosing Spondylitis Functional Index (BASFI) were the outcomes in the current study. The BASDAI is measuring AS specific disease activity in the week prior to completion of the questionnaire by means of six questions regarding 1) fatigue/tiredness, 2) AS related neck, back or hip pain, 3) pain/swelling in joints other than neck, back or hips, 4) the level of discomfort from an area tender to touch or pressure, 5) the level of morning stiffness and 6) the duration of morning stiffness. Each question is scored on a scale from 0 (none) to 10 (severe). In case of the last question, 0 equals 0 hours, 5 equals 1 hour and 10 equals two or more hours. BASDAI-scores range from 0 to 10. Higher values represent more active disease. A score ≥4/10 indicates “active” disease. The final BASDAI score is calculated as follows:

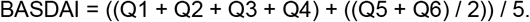

The BASFI is measuring the AS specific degree of functional limitation in the week prior to completion of the questionnaire by means of 10 questions regarding 1) putting on socks, 2) bending forward, 3) reaching up, 4) getting up from a chair, 5) getting up off the floor, 6) standing unsup<u>p</u>orted for 10 minutes without discomfort, 7) climbing steps, 8) looking over shoulder, 9) doing physically demanding activities and 10) doing a full day’s activities at home or at work. Each question is scored on a scale from 0 (easy) to 10 (impossible). The final BASFI score is calculated as follows:

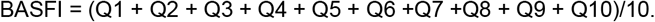

The main independent variable of interest was the timepoint of survey completion distinguishing between timepoint 0 to timepoint 4 (i.e. directly before; directly after; 3; 6; and 9 months after intervention). Covariates were determined a priori due to their already established association with the outcomes and included age, body mass index (BMI) and sex.

### Statistical Analyses

The study sample was characterized in terms of age, BMI and sex at timepoint 0 and in terms of BASDAI and BASFI score at each timepoint using descriptive statistics adequate for the metric properties of each variable. A mixed-effects linear regression model was employed to explore the association between timepoint of measurement (using timepoint 0 as a reference) and the BASDAI and BASFI scores, respectively. Age, sex, and BMI were incorporated as fixed factors in the model. Patient ID was included as a random factor to account for individual variations between patients in terms of unmeasured characteristics and treatment combinations. After each model, the Stata command “margins” was used to produce age, sex and BMI standardized estimates and their 95% confidence interval (CI) for BASDAI and BASFI at each timepoint. P-values ≤0.05 were considered statistically significant. A change of 17.5% in BASFI-scores and of 22.5% in BASDAI-scores was considered clinically relevant.

## Results

The study sample consisted of 249 subjects of whom the average BMI was 26.64 (SD 4.27).

### Alterations in functional impairment

Compared to T0, age, sex and BMI standardized BASFI scores improved significantly until 6 months after treatment. Older age and higher BMI were adversely associated with BASFI scores (Table 1). Based on the fixed portion of the mixed-effects linear regression model, predicted BASFI scores at T0, T1, T2 and T3 were 2.99 (95% CI 2.77; 3.21), 2.31 (95% CI 2.08; 2.53), 2.38 (95% CI 2.15; 2.61) and 2.69 (95% CI 2.46; 2.92). At T4 the score was higher than at T0 (3.04 (95%CI 2.81; 3.28). Compared to T0, the BASFI scores at T1 and T2 reflected a clinically relevant change of approximately 23% and 20%, respectively (Figure 1).

**Table 1:** Alterations in disease activity and functional impairments in ankylosing spondylitis patients after multimodal spa therapy.

|  | BASDAI | BASFI |
| --- | --- | --- |
| | $\beta^*$ [95% Confidence Interval] | |
| TIMEPOINT** |  |  |
| Directly after treatment | **-1.38 [-1.58; -1.18]** | **-0.69 [-0.85; -0.53]** |
| 3 months after treatment | **-1.26 [-1.47; -1.06]** | **-0.61 [-0.78; -0.44]** |
| 6 months after treatment | **-0.86 [-1.06; -0.65]** | **-0.30 [-0.47; -0.13]** |
| 9 months after treatment | **-0.51 [-0.71; -0.30]** | 0.05 [-0.12; 0.22] |
| Age | **0.02 [0.00; 0.04]** | **0.05 [0.03; 0.07]** |
| Sex | 0.24 [-0.12; 0.60] | -0.20 [-0.59; 0.20] |
| Body mass index | 0.01 [-0.03; 0.05] | **0.09 [0.05; 0.14]** |

Results of fixed portion of multivariable mixed-effects models, random factor: patient-ID
\* estimated change in outcome variable for one unit change in independent variable
\*\* reference category: before treatment
Significant p-value: $\leq 0.05$ (bold)

**Figure 1.**
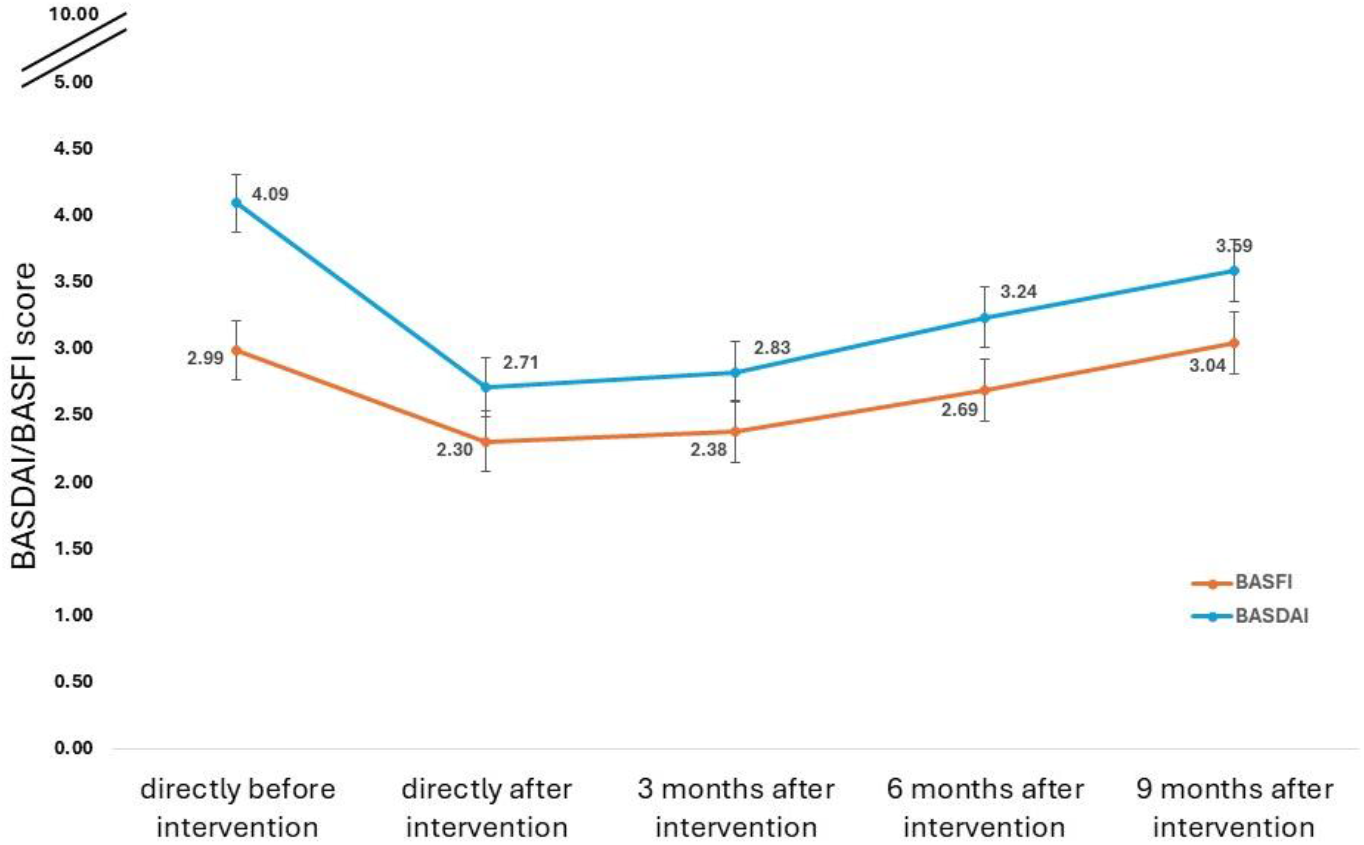
Predicted* margins and 95% confidence intervals of functional impairment (BASFI) and disease activity (BASDAI) in ankylosing spondylitis patients following multimodal spa therapy *Predictions based on fixed portion of multivariable linear mixed models adjusted for age, sex and body mass index, higher scores reflect higher disease activity and more impairments in functional ability

### Alterations in disease activity

Compared to T0, age, sex and BMI standardized BASDAI scores improved significantly until 9 months after treatment. Older age and higher BMI corresponded adversely with BASDAI scores (Table 1). The predicted BASDAI scores at T0, T1, T2, T3 and T4 were 4.09 (95% CI 3.88; 4.31), 2.71 (2.49; 2.94), 2.83 (95% CI 2.60; 3.06), 3.24 (95% CI 3.01; 3.46) and 3.59 (95% CI 3.36; 3.82), respectively. Compared to T0, the BASDAI scores T1, T2 and T3 reflected a clinically relevant change of approximately 34%, 31% and 21%, respectively (Figure 1).

## Discussion

In line with the hypothesis, we found significant improvements in BASFI and BASDAI scores up to 9 months after treatment and clinically relevant improvements until 6 months after treatment for the BASFI and until 9 months after treatment for the BASDAI, while adjusting for age, sex and BMI and accounting for unmeasured individual differences in patient characteristics.

In accordance with the findings of several previous studies, the present research underscores the benefits of spa therapy in combination with low-dose radon in enhancing function and disease activity in AS patients [14, 17–20]. For instance, a randomized controlled trial (RCT) by van Tubergen et al. showed that a combined spa-exercise therapy incorporating low-dose radon speleotherapy compared to standard pharmacological treatment in patients with AS significantly improved both functional performance and disease activity up to 40 weeks after treatment [20]. With regard to the point in time at which the greatest improvement is observed, there is a very heterogeneous body of evidence. The RCT by van Tubergen et al. demonstrated the most pronounced improvements in function and disease activity at 16 weeks after treatment, in accordance with findings of a prospective study which showed the greatest enhancements in terms of functional capacity, disease activity, and pain at 3 months post-treatment [14, 20]. In contrast, the present study suggests that the greatest improvements are evident immediately after spa therapy incorporating low-dose radon balneo- and/or speleotherapy. These immediate improvements are consistent with the findings of a pilot study that examined the efficacy of low-dose radon and hyperthermia treatment, comprising ten visits to a radon healing gallery over a 3-week period, which found significant improvements in BASDAI and BASFI scores immediately after treatment [18]. The heterogeneity in treatment outcomes might be attributed to differences in the patients’ baseline characteristics. For example, patients with less inflammation and impairments in function might respond differently than those with more severe symptoms. In the view of the absence of this information in the majority of the studies that investigated the topic, comparisons of improvement differences must be undertaken with caution. Like with the body of literature addressing the point of greatest improvement, there is heterogeneity with regard to the duration of the post-treatment effect in terms of BASFI and BASDAI. A substantial discrepancy in the design of the studies was observed, with a study duration ranging from no follow-up to 6 months following treatment [14, 18–20]. Consequently, a direct comparison between the studies is not without challenges. However, while the present study and the RCT by van Tubergen et al. indicate a long-term effect in the enhancement of BASFI and BASDAI, the study by Moder et al. has been limited to assessment at baseline and end of therapy, leaving long-term effects unclear and emphasizing the necessity for more longitudinal studies [18, 20].

In a small sample of AS patients, Franke et al. found improved (although insignificantly) BASFI scores [17]. These scores were part of a larger set of outcomes focusing on patient well-being after low-dose radon treatment. The integration of functional endpoints and outcomes focusing on disease activity into questionnaire batteries that address the multidimensionality of health is linked with the increasing awareness for individual patient perspectives with regard to their health and the relevance of such outcomes in the context of research [17]. This findings align with the recent EULAR recommendations, emphasizing the importance of QoL in this context [6]. A longitudinal analysis of prospectively collected registry data investigating the effect of multimodal spa therapy, including low-dose radon, on QoL scores and pain intensity has yielded significantly higher EQ-5D-5L scores. The findings indicate clinically significant improvements in pain scores and QoL utilities, including functional mobility, up to 9 months after treatment, thereby underscoring the aforementioned recommendations [15]. In the context of studies examining multimodal therapy in isolation, the findings indicate that this holistic approach can complement pharmacological treatment. A systematic review investigating the effects of multidisciplinary non-pharmacological treatments on pain and functioning in individuals with AS found multiple methods of treating AS, primarily focused on balneo- and physiotherapy. The review’s findings indicate promising effects of those complementary approaches [12]. In consideration of the importance of spa therapy comprising low-dose radon for individuals diagnosed with AS, the present study posits that this special treatment modality, in conjunction with a multimodal therapeutic framework, holds considerable significance within the broader spectrum of non-pharmacological treatment strategies.

However, it is important to note that all studies that are derived from registry data are constrained by inherent limitations. This is primarily due to the fact that data collection is not directly monitored or conducted by the researcher. In addition, the accessibility of data pertaining to confounders is frequently restricted [21, 22]. In relation to the present study, a potential limitation is that data concerning the frequency of interventions prior to the initial measurement point were not systematically collected. The collection of such data might have provided a more accurate picture of the baseline values, as participants who have received the intervention repeatedly over time likely have a better baseline health state than those who receive the intervention for the first time. This possible bias might cause the improvement observed in first-time participants to be underestimated. Additionally, it is crucial to recognize that in pre-post evaluations, self-reported measures can be influenced by the intervention itself, potentially affecting participants’ perception of their health rather than reflecting true changes in their health status. The study’s strengths include a relatively large sample size and real-world data over an extended period, thereby minimizing the effects of selection and attrition bias. Moreover, the independence of data collection ensures the integrity of the results [22]. In addition, registries represent a cost-effective approach in comparison to other research designs and facilitate the description of disease outcomes and outcome patterns over an extended period of time [21]. In light of the paucity of data concerning the effectiveness of spa therapy, including radon, on the improvement of functional performance and disease activity in AS patients, the present study provides significant insights and opportunities for further research, including comparative studies with usual care and among other patient populations.

In conclusion, the present study shows that AS patients undergoing multimodal spa therapy comprising low-dose radon exhibit substantial and clinically significant improvements in their functional performance and disease activity. These effects persist for a period of up to 9 months. Hence, this treatment modality may serve as a valuable complementary therapeutic option for patients with AS. The findings of this study may provide a rationale for policymakers and insurance providers to consider reimbursement for biannual spa therapy incorporating radon for patients with AS.

## Data Availability

All data produced in the present study are available upon reasonable request to the authors

## Abbreviations

ASAS: Assessment of Spondyloarthritis international Society
BASDAI: Bath Ankylosing Spondylitis Disease Activity Index
BASFI: Bath Ankylosing Spondylitis Functional Index
BMI: body mass index
CI: confidence interval
EULAR: European League Against Rheumatism
JAK: Janus kinase
NSAID: nonsteroidal anti-inflammatory drug
PRO: patient-reported outcome
QoL: quality of life
RCT: randomized controlled trial
RMD: rheumatic musculoskeletal disorder
SpA: spondyloarthropathy
TNFi: TNF-α inhibitor

## Ethics approval

The studies involving human participants were reviewed and approved by Ethics Committee of the County of Salzburg (Amt der Salzburger Landesregierung, Ethikkommission für das Bundesland Salzburg, Postfach 527, 5010 Salzburg (No. 415– E/1966/3–2015)). The patients/participants provided their written informed consent to participate in this study.

## Acknowledgements

We would like to thank all investigators, study personnel and participants who have contributed to the “Radon indication registry”. We would like to acknowledge, that the findings of the study were presented at the EULAR conference 2024. (van der Zee-Neuen A, Bogensberger K, Fuchs J, Wildburger S, Ritter M. AB0937 - IMPROVEMENTS IN DISEASE ACTIVITY AND FUNCTIONAL IMPAIRMENT AFTER MULTIMODAL SPA THERAPY IN ANKYLOSING SPONDYLITIS PATIENTS. ANNALS OF THE RHEUMATIC DISEASES. 2024;2024(83):1778. doi: 10.1136/annrheumdis-2024-eular.827)

## Author Contributions

SW, JF, MG, MR, and AvZN conceived the project. BH, RR, WF, HR, and MO were local study coordinators at the participating facilities. AvZN planned the study and developed the research design. JF prepared and organized the data before analysis. AvZN performed the analyses and SW, JF, KB, and AvZN discussed and interpreted the data. KB and AvZN drafted the article, revised it critically and approved the final version for publication. SW, JF, BH, RR, WF, HR, MO, MG, and MR reviewed and critically commented on the manuscript. All authors approved the final manuscript.

## Patient and public involvement statement

No patients were involved in the development of the study design, conduct of the study or reporting of the data.

## References

1. Zhu W, He X, Cheng K, et al. Ankylosing spondylitis: etiology, pathogenesis, and treatments. Bone Res. 2019;7:22.

2. Moltó A, Nikiphorou E. Comorbidities in Spondyloarthritis. Front Med (Lausanne). 2018;5:62.

3. Lindström U, Olofsson T, Wedrén S, et al. Impact of extra-articular spondyloarthritis manifestations and comorbidities on drug retention of a first TNF-inhibitor in ankylosing spondylitis: a population-based nationwide study. RMD Open. 2018;4(2):e000762.

4. Braun J, Sieper J. Ankylosing spondylitis. Lancet. 2007;369(9570):1379–90.

5. Mauro D, Thomas R, Guggino G, et al. Ankylosing spondylitis: an autoimmune or autoinflammatory disease? Nature Reviews Rheumatology. 2021;17(7):387–404.

6. Ramiro S, Nikiphorou E, Sepriano A, et al. ASAS-EULAR recommendations for the management of axial spondyloarthritis: 2022 update. Ann Rheum Dis. 2023;82(1):19–34.

7. Danve A, Deodhar A. Treatment of axial spondyloarthritis: an update. Nat Rev Rheumatol. 2022;18(4):205–16.

8. Reimold AM, Chandran V. Nonpharmacologic therapies in spondyloarthritis. Best Pract Res Clin Rheumatol. 2014;28(5):779–92.

9. Kocyigit BF, Sagtaganov Z, Yessirkepov M, et al. Assessment of complementary and alternative medicine methods in the management of ankylosing spondylitis, rheumatoid arthritis, and fibromyalgia syndrome. Rheumatol Int. 2023;43(4):617–25.

10. Packham J. Optimizing outcomes for ankylosing spondylitis and axial spondyloarthritis patients: a holistic approach to care. Rheumatology (Oxford). 2018;57(suppl_6):vi29–vi34.

11. Jeziorski K. Hyperthermia in rheumatic diseases. A promising approach? Reumatologia. 2018;56(5):316–20.

12. Ungureanu A-E, Stanciu L, Uzun A-B, et al. Multidisciplinary non-pharmacological treatments with effectson pain modulation and functioning in spondyloarthropathies– a systematic review. 2023.

13. Bekaryssova D, Yessirkepov M, Imanbaeva AD. Water-based interventions in rheumatic diseases: mechanisms, benefits, and clinical applications. Rheumatol Int. 2024;45(1):8.

14. Dischereit G, Neumann N, Müller-Ladner U, et al. Einfluss einer seriellen niedrig-dosierten Radonstollen-Hyperthermie auf Schmerz, Krankheitsaktivität und zentrale Zytokine des Knochenmetabolismus bei ankylosierender Spondylitis – eine Prospektivstudie. Aktuelle Rheumatologie. 2014;39:304–9.

15. van der Zee-Neuen A, Strobl V, Dobias H, et al. Sustained improvements in EQ-5D utility scores and self-rated health status in patients with ankylosing spondylitis after spa treatment including low-dose radon – an analysis of prospective radon indication registry data. BMC Musculoskeletal Disorders. 2022;23.

16. Kiltz U, Braun J. Assessments of Functioning in Patients With Axial Spondyloarthritis. jrd. 2019;27(1):22–9.

17. Franke A, Reiner L, Resch KL. Long-term benefit of radon spa therapy in the rehabilitation of rheumatoid arthritis: a randomised, double-blinded trial. Rheumatol Int. 2007;27(8):703–13.

18. Moder, Hufnagl A, Lind-Albrecht C, et al. Effect of combined Low-Dose Radon- and Hyperthermia Treatment of Patients with Ankylosing Spondylitis on Serum Levels of Cytokines and Bone Metabolism Markers: A Pilot Study. International Journal of Low Radiation. 2010;7.

19. Moder A, Hufnagl C, Jakab M, et al. Radon-Therapy in Ankylosing Spondylitis reduces auto-Antibody titers. Open Journal of Molecular and Integrative Physiology. 2011;1:52–4.

20. van Tubergen A, Landewé R, van der Heijde D, et al. Combined spa-exercise therapy is effective in patients with ankylosing spondylitis: a randomized controlled trial. Arthritis Rheum. 2001;45(5):430–8.

21. Psoter KJ, Rosenfeld M. Opportunities and pitfalls of registry data for clinical research. Paediatr Respir Rev. 2013;14(3):141–5.

22. Thygesen L, Ersbøll A. When the entire population is the sample: Strengths and limitations in register-based epidemiology. European journal of epidemiology. 2014;29.

